# Effect of omega-3 LCPUFA supplementation on maternal fatty acid and oxylipin concentrations during pregnancy

**DOI:** 10.1101/2020.06.22.20137851

**Authors:** K P Best, R A Gibson, L N Yelland, S Leemaqz, J Gomersall, G Liu, M Makrides

## Abstract

**Introduction:** Omega-3 long chain polyunsaturated fatty acids (LCPUFA) have been associated with a reduction in risk for preterm birth. However, there is limited understanding of how fatty acids and their bioactive derivatives (oxylipins) change over the course of pregnancy. Here we document the changes in concentration of fatty acids and oxylipins during pregnancy and how fatty acid status and oxylipin concentrations are affected by supplementation with omega-3 LCPUFA. We also investigate the degree to which fatty acid and oxylipin changes across pregnancy are influenced by baseline omega-3 status.

**Materials and Methods:** We profiled the fatty acids in all lipids in dried blood spots (total blood fatty acids) by gas chromatography and free (unesterified) fatty acids and their associated oxylipins in separate dried blood spot samples by LC-MS-MS collected from a random sample of 1263 women with a singleton pregnancy who participated in the ORIP (Omega-3 fats to Reduce the Incidence of Prematurity) trial. ORIP is a double-blind, randomized controlled trial involving 5544 participants and designed to determine the effect of supplementing the diets of pregnant women with omega-3 LCPUFA on the incidence of early preterm birth. Maternal whole blood finger prick samples were collected at baseline (∼14 weeks gestation) and at completion of the study intervention period (34 weeks gestation).

**Results:** The concentration of most total and free polyunsaturated fatty acids and their associated oxylipins declined over the course of pregnancy. Omega-3 LCPUFA supplementation increased total DHA and 7-HDHA and mitigated the decline in free DHA, 4-HDHA and 14-HDHA. The intervention had minimal or no effect on free EPA, LA, AA and their associated oxylipins. Omega-3 LCPUFA supplementation in women with higher omega-3 status at baseline was associated with a significant increase in 7-HDHA and 4-HDHA between the treatment and control whereas there were no differences between groups in 7-HDHA and 4-HDHA in women with intermediate or lower baseline omega-3 status.

**Conclusion:** Our data suggest a differential response with or without omega-3 supplementation for DHA and DHA-derived oxylipins, which may have an important role to play in modulating pregnancy duration. Further work is needed to understand their role, which may allow us to better tailor omega-3 supplementation for preterm birth prevention.

## 1. Introduction

There is strong evidence to support maternal supplementation with omega-3 long chain polyunsaturated fatty acids (LCPUFAs) to reduce the risk of preterm birth [1, 2]. Some of the pro- or anti-inflammatory effects of LCPUFAs can be attributed to their oxidized products, collectively known as oxylipins. Oxylipins comprise a large, heterogeneous family of bioactive mediators that have essential roles in normal physiology and function [3]. Synthesis usually occurs via the cyclooxygenase, lipoxygenase or cytochrome P450 pathways [4] and alterations in oxylipin concentrations have been described for numerous diseases in human and animal models [5-9].

During pregnancy, oxylipins are known to alter the function of the placenta, vascular and immune responses [9-11], yet little is known about how total and free fatty acid and oxylipin levels change during pregnancy. Our recent secondary analysis of singleton pregnancies in the ‘Omega-3 fats to Reduce the Incidence of Prematurity’ (ORIP) trial, suggests that the sum of all omega-3 fatty acids (docosahexaenoic acid [DHA] + eicosapentaenoic acid [EPA] + docosapentaenoic acid [DPA] + alpha linolenic acid [ALA]) in whole blood lipids in early pregnancy, is the strongest biomarker to predict early preterm birth. We found that women with a low omega-3 baseline status (≤4.1% of total fatty acids) are at higher risk of early preterm birth (<34 weeks gestation) and that this risk can be reduced by omega-3 LCPUFA supplementation. However, the data also suggest that supplementing mothers who are already replete in omega-3 (>4.9% total fatty acids) with additional omega-3 LCPUFA, significantly increases the risk of early preterm birth [12]. Because of the high biological activity of LCPUFAs and their derived oxylipins [13-15], we sought to determine whether omega-3 LCPUFA supplementation leads to oxylipin changes that may be critical to prevent preterm birth.

Using maternal fatty acid status data from blood collected from singleton pregnancies at entry to our ORIP trial (∼14 weeks’ gestation), we explored the changes in concentration of whole blood lipid fatty acids, free fatty acids and oxylipins during pregnancy from around 14 to 34 weeks’ gestation. Here we describe differences between women who were supplemented with omega-3 LCPUFA and non-supplemented women in the control group. Because the ORIP Trial intervention contained 90% DHA and 10% EPA, our analyses focused on these omega-3 LCPUFA. Since it is possible that some interconversion of omega 3 fatty acids occurred, we examined changes in whole blood lipid omega-3 status (sum of all omega-3 fatty acids; DHA, EPA, DPA and ALA) and free omega-3 fatty acids. Key competitive omega-6 fatty acids, arachidonic acid (AA) and linolenic acid (LA) were also examined. In particular, to explore differential effects of omega-3 LCPUFA supplementation in pregnancy [12], we also report the degree to which the baseline omega-3 status altered the changes in the concentration of free fatty acids and oxylipins over time with treatment.

## 2. Participants and Methods

### 2.1. ORIP Trial - Sample Collection

The ORIP trial was a multicenter, double-blind, randomized controlled trial designed to determine the effect of supplementing the diets of pregnant women with omega-3 LCPUFA on the incidence of early preterm birth [16, 17]. The ORIP trial primary results have been previously published [17]. Women were recruited to the ORIP trial prior to 20 weeks’ gestation, with few exclusions, including those who had been regularly consuming a low dose DHA supplement (≤150 mg per day) and were willing to cease. A total of 5544 women were randomized to receive either ≈900 mg omega-3 LCPUFA (3 x DHA enriched fish oil/day (≈800 mg DHA and ≈100 mg eicosapentaenoic acid (EPA)/day)) or isocaloric vegetable oil control capsules with trace fish oil for masking (≈15 mg DHA and ≈4 mg EPA/day) until 34 weeks of gestation. A finger prick blood sample was collected from participants on to chemically treated filter paper as two dried blood spots [18] at baseline (∼14 weeks’) and 34 weeks’ gestation [17]. A simple random sample of 1382 ORIP women were selected for further detailed analyses.

The present study reports findings from additional analyses of dried blood spot samples to measure concentration of fatty acids and their associated oxylipins among women in this random sample who had a singleton pregnancy. Measures in maternal capillary whole blood, collected at ∼14 and 34 weeks’ gestation, are reported in **Supplementary Table 1**.

### 2.2. Laboratory Analysis

All fatty acid analyses were undertaken in the fatty acid laboratory at the School of Agriculture and Wine, University of Adelaide. Briefly, the dried blood spot on a chemically treated paper (18) was treated as follows:

For total fatty acids (the complete fatty acid spectrum from all lipids in blood), the first blood spot was placed in a 6mL vial (Wheaton, Millville, USA) with 2 mL of 1% (v/v) H2SO4 (18M, AR grade, BDH, Sussex, UK) in anhydrous methanol (Merck, Darmstadt, Germany), sealed and heated at 70°C for 3 hours. The resultant fatty acid methyl esters (FAME) were extracted into heptane (Merck, Darmstadt, Germany). FAME were separated and quantified using an Agilent 7890 Gas Chromatograph (Agilent, CA, USA) equipped with a BPX70 capillary column 30 m x 0.25 mm, film thickness 0.25 µm (Trajan Pty Ltd., Victoria, Australia), programmed temperature vaporization injector (at 250°C) and a flame ionization detector (at 300°C). A programmed temperature ramp (140-240°C) was used. Helium gas was utilized as a carrier at a flow rate of 1 mL/min in the column and the inlet split ratio was set at 20:1. The FAME were identified and quantified by comparing the retention times and peak area values of unknown samples to those of commercial lipid standards (Nu-Chek Prep Inc., Elysian, MN, USA) using the Agilent Chemstation data system.

For free fatty acids (non-esterifed fatty acids) and their oxylipins, a 6 mm disc of blood was obtained from the second dried blood spot and placed into a separated well in a 96-well plate with extraction solvent (150 µL of 80% aqueous methanol) containing deuterated internal standards (d5-DHA, d8-AA, d5-EPA, d4-LA, d8-12S-HETE, d4-LTB4 and d4-13S-HODE). The plate was covered and gently shaken on a plate shaker for 30 min at room temperature, and the extract was analyzed using an Agilent 1290 infinity ultra-high-performance liquid chromatography equipped with AB SCIEX 5500 triple quadrupole tandem mass spectrometer, using [electrospray ionisation](https://www-sciencedirect-com.proxy.library.adelaide.edu.au/topics/medicine-and-dentistry/electrospray-ionization) in negative mode. The mass [spectrometer](https://www-sciencedirect-com.proxy.library.adelaide.edu.au/topics/medicine-and-dentistry/spectrometer) had the following conditions: nitrogen was used as the source gas; curtain gas and collision gas were set at 20 and 6 arbitrary units, respectively; capillary voltage was −4500 V; entrance potential was set at −10 V and temperature was set at 200 °C [19, 20].

### 2.3. Statistical Analysis

A separate analysis was performed for each fatty acid and oxylipin. The analysis was performed on the available data and any values below the limit of detection were set to half the limit of detection. The data were analyzed using a linear model including treatment group, time point and their interaction, with adjustment for the randomization strata (enrolment center and recent omega-3 supplementation). Clustering due to repeated measurements over time was taken into account using generalized estimating equations with an exchangeable working correlation structure. Post hoc comparisons were performed to test whether there were changes in the concentration of the fatty acids and oxylipins over time (34 weeks’ vs baseline) separately within each group, and whether there were differences between the groups in the changes in concentration over time (omega-3 change over time vs control change over time). Statistical significance was assessed at the P<0.05 level and 95% confidence intervals were calculated for all estimated effects, without adjustment for multiple comparisons.

A sensitivity analysis was performed using multilevel tobit regression models, where values below the limit of detection were treated as censored. The multilevel tobit regression approach was chosen to be a sensitivity analysis, rather than the primary analysis, due to the small number of oxylipins affected, the generally small percentage of values below the limit of detection, and concerns over the constant variance assumption required for the multilevel tobit regression model.

Exploratory analyses were performed for DHA and total omega-3 fatty acids to investigate whether the changes in the concentration of fatty acids and oxylipins over time within each treatment group and the differences between treatment groups depended on baseline omega-3 status. Women were categorized as having low (≤4.1%), moderate (>4.1 to ≤4.9%) or high (>4.9%) baseline omega-3 status, corresponding to subgroups where omega-3 LCPUFA supplementation has been shown to significantly reduce the risk, have no significant effect, or significantly increase the risk of early preterm birth, respectively [12]. The analysis model included treatment group, time point, baseline omega-3 status category and all 2 and 3-way interactions. Post hoc comparisons were performed to test for differences between the high and low status categories (>4.9% vs ≤4.1%) in the change over time within groups and the difference between groups in these changes over time. All analyses were performed using R version 3.6.2.

## 3. Results

### 3.1. Characteristics of the study population

Of the 5544 women enrolled in the ORIP Trial, 1382 women were randomly selected to have their blood samples undergo further analysis to determine fatty acid and oxylipin concentrations. This report presents findings for the 1263 women in this random sample who had a singleton pregnancy and baseline dried blood spot data, of which 1023 (81%) also had a dried blood spot sample at 34 weeks’ gestation, **Figure 1**. Descriptive analyses showed that women in both groups had comparable baseline characteristics and fatty acid profiles as expected due to randomization, **Table 1**. Oxylipins at baseline were also similar by group, as shown in **Supplementary Table 2**.

**Table 1.** Baseline Characteristics of women included in the analysis by treatment group.

| Characteristic | Omega-3 (N=614) | Control (N=649) | Total (n=1263) |
| --- | --- | --- | --- |
| Consumed omega-3 containing supplements in the previous 3 mo: n(%) | 89 (14.5) | 85 (13.1) | 174 (13.8) |
| Maternal age: Median (IQ range) | 30.0 (27.0-34.0) | 30.0 (27.0-34.0) | 30.0 (27.0-34.0) |
| Gestation: Median (IQ range)* | 14.0 (12.6-16.1) | 14.1 (12.7-16.4) | 14.0 (12.7-16.3) |
| Mother was primiparous: n/N(%) | 280/614 (45.8) | 280/648 (43.2) | 560/1260 (44.4) |
| Maternal weight at trial entry: Median (IQ range)** | 69.0 (60.0-81.0) | 69.0 (60.4-81.4) | 69.0 (60.0-81.0) |
| Maternal white race: n/N(%) | 456/614 (74.3) | 456/648 (70.4) | 912/1262 (72.3) |
| Mother completed a high-school education: n/N(%) | 506/612 (82.7) | 499/647 (77.1) | 1005/1259 (79.8) |
| Mother smoked cigarettes at trial entry or leading up to pregnancy: n/N(%) | 95/612 (15.5) | 101/648 (15.6) | 196/1260 (15.6) |
| Mother drank alcohol at trial entry or leading up to pregnancy: n/N(%) | 356/612 (58.2) | 362/648 (55.9) | 718/1260 (57.0) |
| Mother had previous preterm delivery: <37 wk of gestation: n/N(%) | 37/612 (6.0) | 39/648 (6.0) | 76/1260 (6.0) |
| Maternal DHA level (% of total fatty acids): Median (IQ range) | 2.7 (2.3-3.1) | 2.7 (2.3-3.1) | 2.7 (2.3-3.1) |
| Male fetus: n/N(%) | 315/605 (52.1) | 317/636 (49.8) | 632/1241 (50.9) |
| Fatty acid status |  |  |  |
| - Total DHA: Mean (SD) | 2.7 (0.6) | 2.8 (0.7) | 2.7 (0.6) |
| - Free DHA: Mean (SD) | 0.9 (0.5) | 0.9 (0.4) | 0.9 (0.5) |
| - Total EPA: Mean (SD) | 0.5 (0.2) | 0.5 (0.2) | 0.5 (0.2) |
| - Free EPA: Mean (SD) | 0.1 (0.1) | 0.1 (0.1) | 0.1 (0.1) |
| - Omega-3<sup>†</sup>: Mean (SD) | 4.9 (0.8) | 4.9 (1.0) | 4.9 (0.9) |
| - Free Omega-3<sup>†</sup>: Mean (SD) | 1.1 (0.6) | 1.1 (0.5) | 1.1 (0.6) |
| - Total LA: Mean (SD) | 19.4 (2.5) | 19.3 (2.5) | 19.3 (2.5) |
| - Free LA: Mean (SD) | 3.8 (2.5) | 3.8 (2.1) | 3.8 (2.3) |
| - Total AA: Mean (SD) | 8.2 (1.2) | 8.2 (1.2) | 8.2 (1.2) |
| - Free AA: Mean (SD) | 1.2 (0.7) | 1.2 (0.6) | 1.2 (0.6) |

\* Data were missing for 2 pregnancies in the Omega-3 group.
\*\* Data were missing for 11 pregnancies in the Omega-3 group and 19 in the Control group.
<sup>†</sup>Omega-3 fatty acids, sum of docosahexaenoic acid, eicosapentaenoic acid, docosapentaenoic acid and alpha linolenic acid.

**Figure 1.**
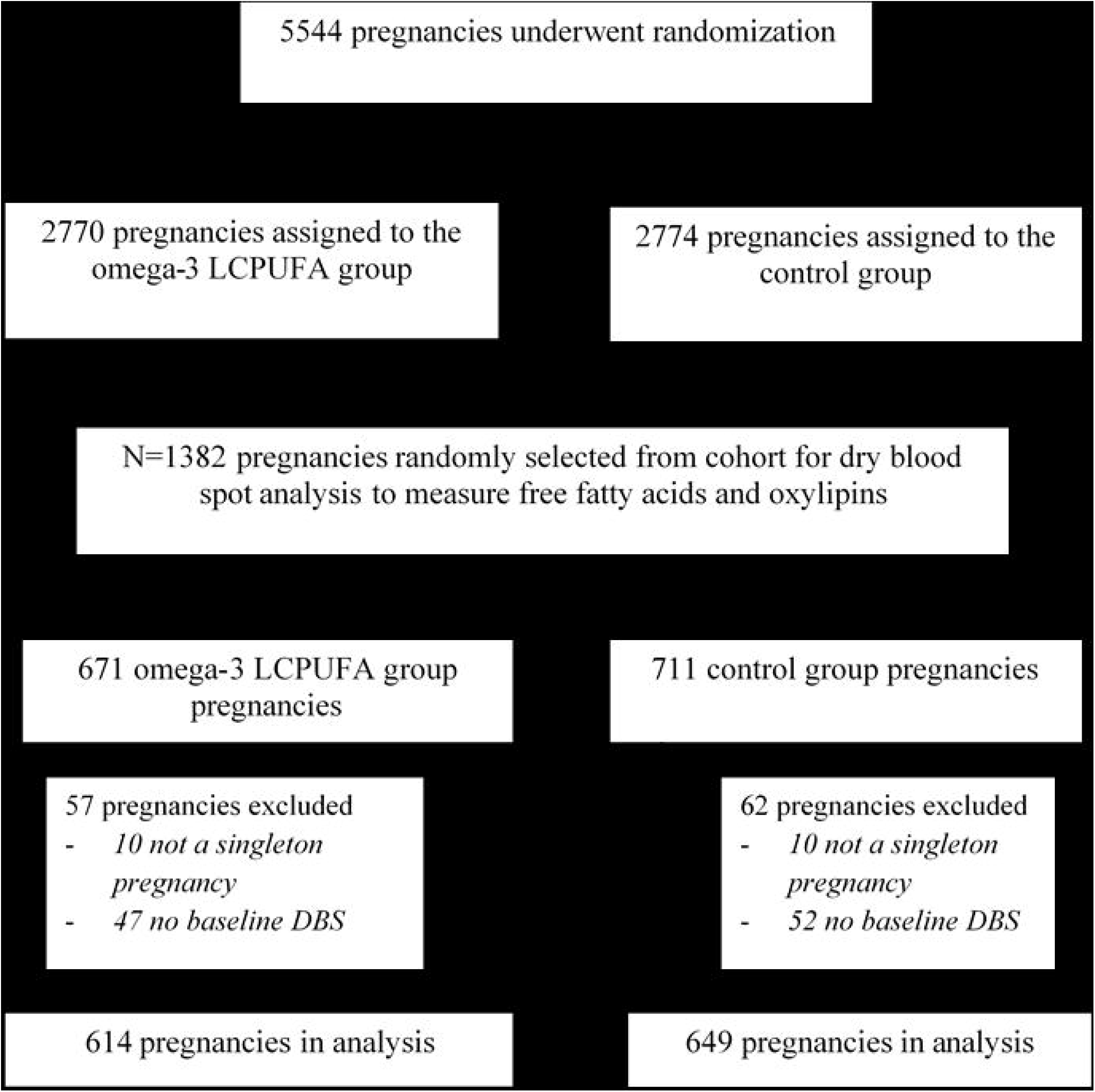
Flowchart of participants in the ORIP trial and the data available for this exploratory analysis. DBS, dried blood spot; LCUPA, long chain polyunsaturated fatty acid; ORIP, Omega-3 fats to Reduce the Incidence of Prematurity

### 3.2. Changes in concentration of fatty acids and oxylipins over time and between groups

The concentration of most fatty acids and oxylipins changed significantly over pregnancy in both the omega-3 LCPUFA and control groups. While the concentration of most fatty acids and oxylipins decreased with gestational age, some increases were observed.

### Omega-3 Fatty Acid Results

#### 3.2.1. DHA

Total DHA decreased during pregnancy in the control group. This was more than offset by supplementation with omega-3 LCPUFA. In contrast, free DHA decreased over pregnancy in both groups, but the drop was significantly smaller in the omega-3 LCPUFA group. This pattern seen for free DHA was mirrored for oxylipins 4-HDHA and 14-HDHA, while 7-HDHA followed the same pattern as total DHA. Omega-3 intervention had no significant effect on the change over pregnancy in 19,20 EpDPA, **Figure 2**.

**Figure 2.**
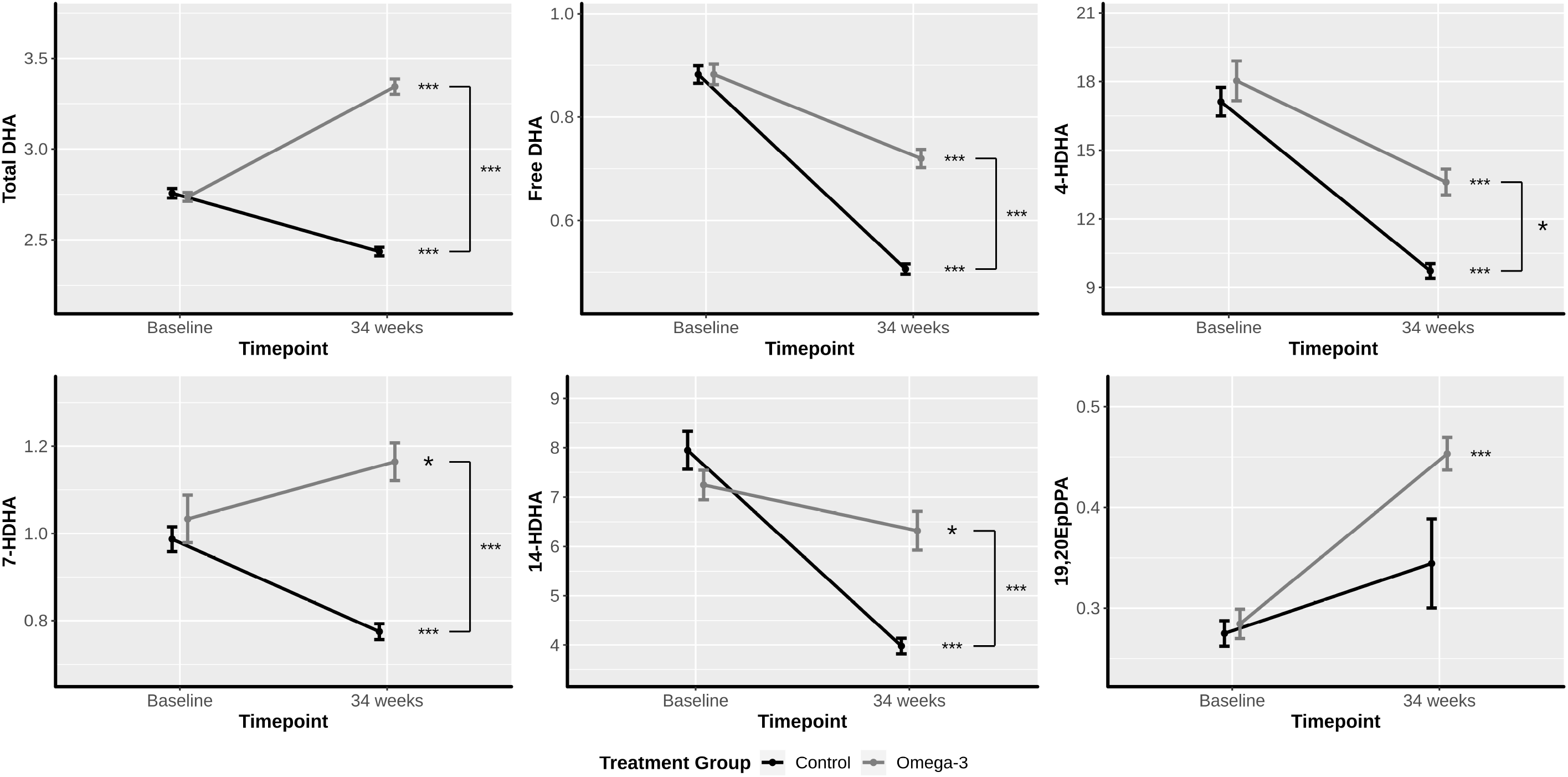
Effect of treatment group and time on concentration of DHA and associated oxylipins. DHA, docosahexaenoic acid Data points are mean fatty acid and oxylipin concentrations by treatment group and time point and error bars are standard errors. Statistical significance of changes over time within treatment groups and differences between treatment groups in the changes over time is indicated by * P<0.05, ** P<0.01 and *** P<0.0001.

#### 3.2.2. EPA

Total EPA decreased significantly over the course of pregnancy, and this was largely ablated by the omega-3 LCPUFA intervention. The changes in free EPA during pregnancy largely reflected those seen in the total EPA with the omega-3 intervention maintaining baseline levels. There was no effect of omega-3 LCPUFA supplementation on the EPA derived oxylipin 18-HEPE, with both groups showing a significant decline over pregnancy **Figure 3**.

**Figure 3.**
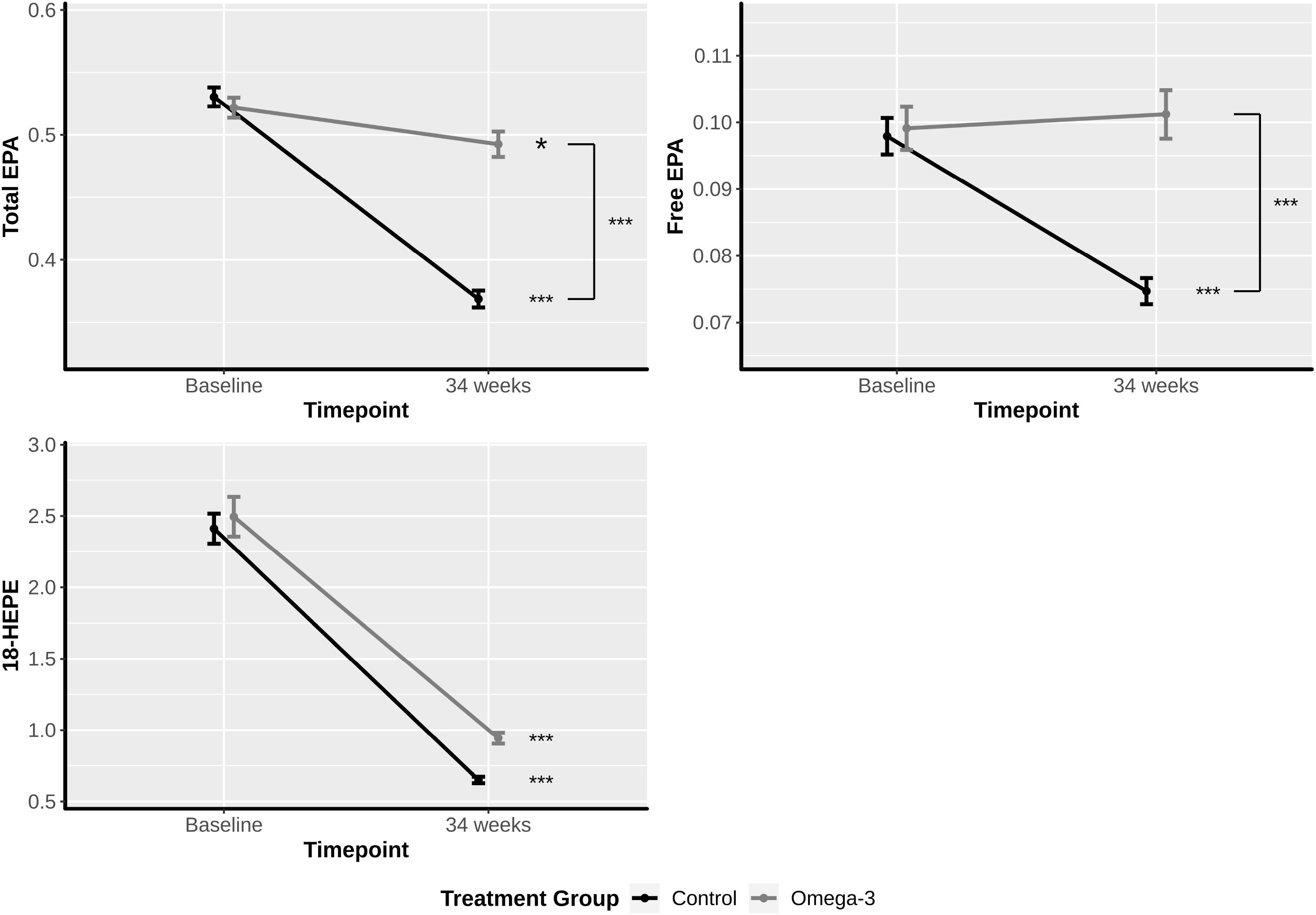
Effect of treatment group and time on concentration of EPA and associated oxylipins. EPA, eicosapentaenoic acid. Data points are mean fatty acid and oxylipin concentrations by treatment group and time point and error bars are standard errors. Statistical significance of changes over time within treatment groups and differences between treatment groups in the changes over time is indicated by * P<0.05, ** P<0.01 and *** P<0.0001.

#### 3.2.3. Omega-3 Fatty Acids Status

The sum of the omega-3 fatty acids in the lipids in dried blood spot (ALA, EPA, DPA and DHA) decreased in the control group from baseline to 34 weeks but increased in the omega-3 LCPUFA supplemented group. The aggregate of free omega-3 fatty acids declined with gestational age in both groups, though the decline was larger in the control group **Figure 6**.

**Figure 4.**
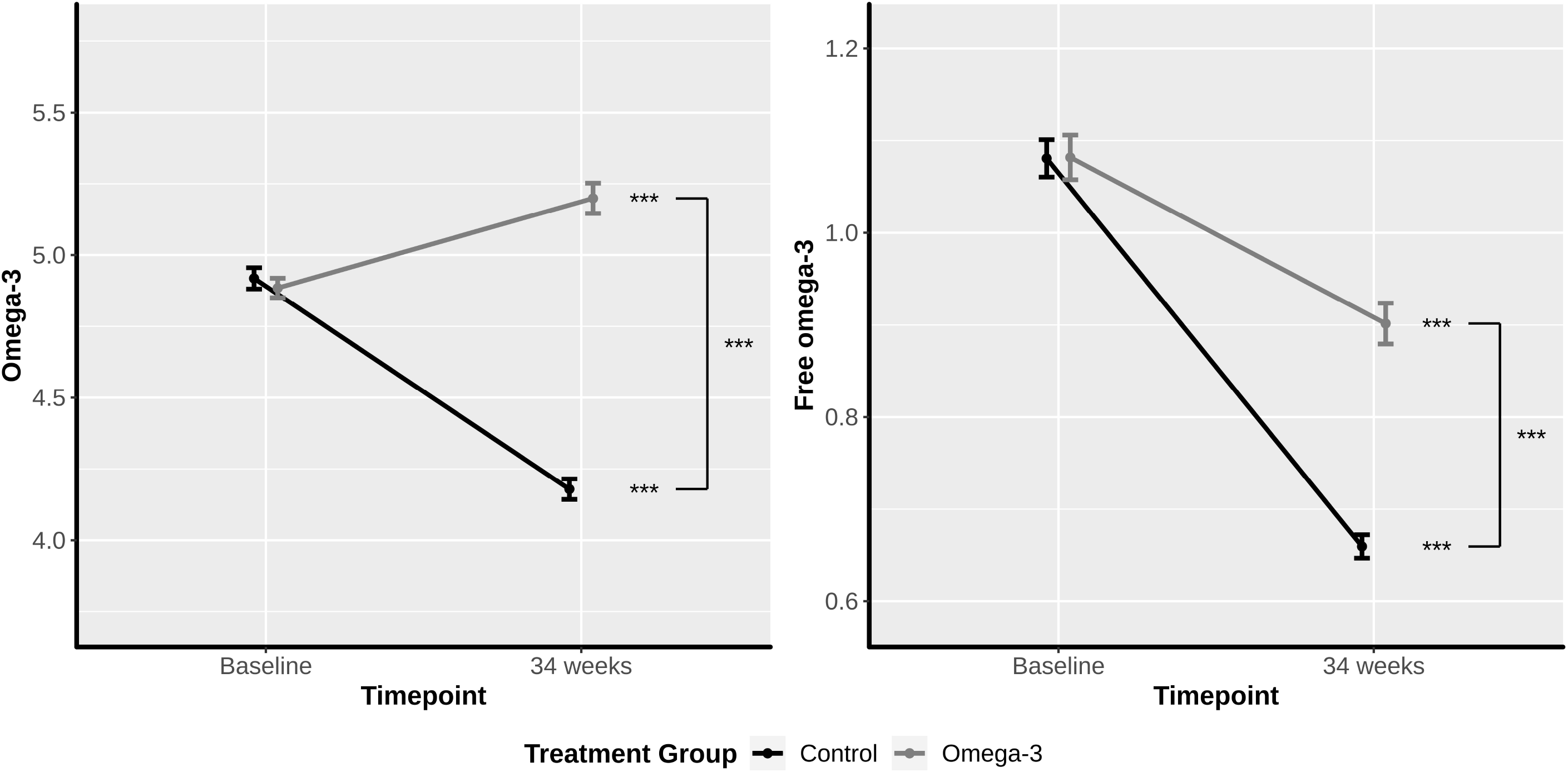
Effect of treatment group and time on concentration of Omega-3. ‘Omega-3’, sum of docosahexaenoic acid, eicosapentaenoic acid, docosapentaenoic acid and alpha linolenic acid. Data points are mean fatty acid and oxylipin concentrations by treatment group and time point and error bars are standard errors. Statistical significance of changes over time within treatment groups and differences between treatment groups in the changes over time is indicated by * P<0.05, ** P<0.01 and *** P<0.0001.

**Figure 5.**
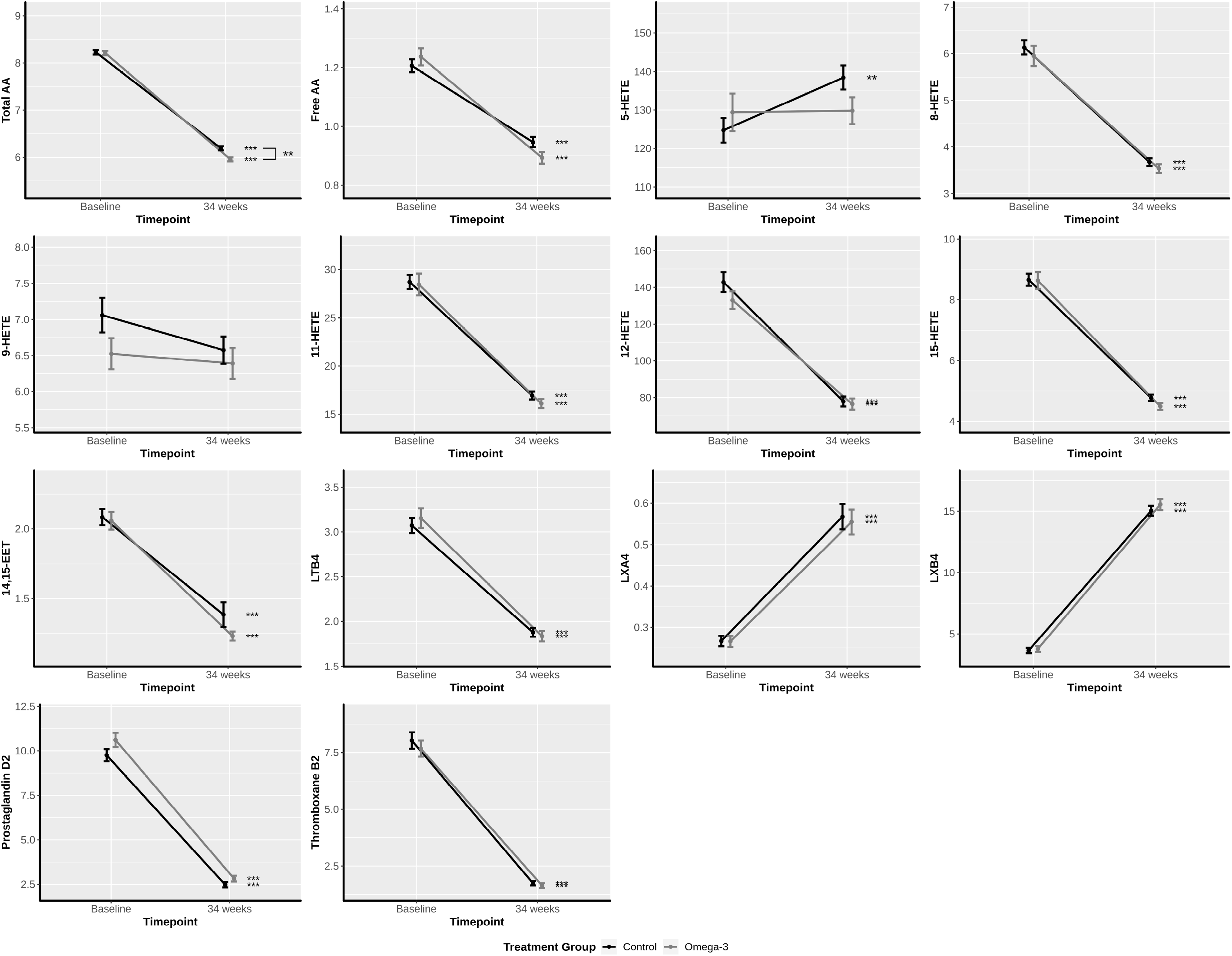
Effect of treatment group and time on concentration of AA and associated oxylipins. AA, Arachidonic acid. Data points are mean fatty acid and oxylipin concentrations by treatment group and time point and error bars are standard errors. Statistical significance of changes over time within treatment groups and differences between treatment groups in the changes over time is indicated by * P<0.05, ** P<0.01 and *** P<0.0001.

**Figure 6.**
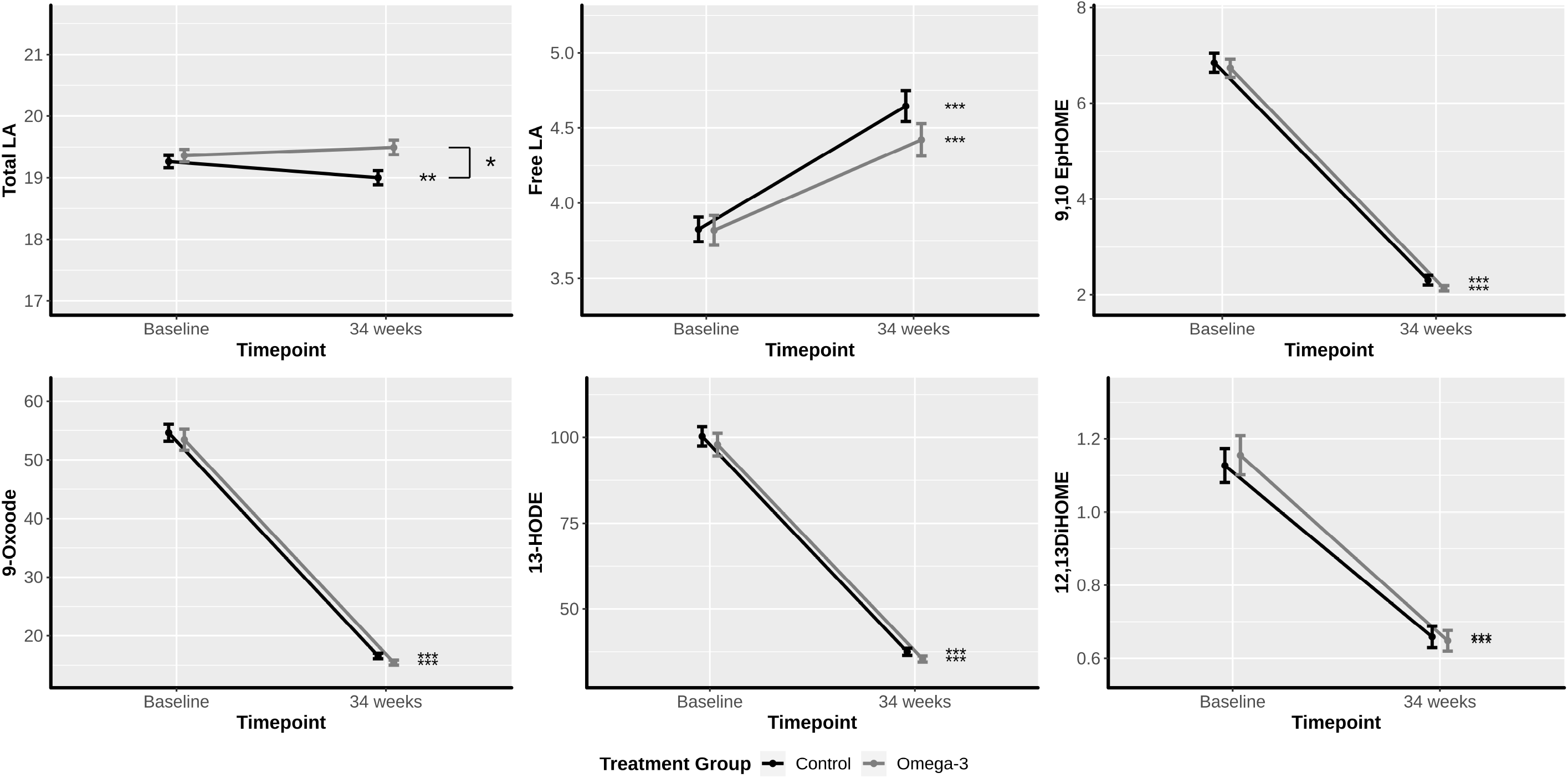
Effect of treatment group and time on concentration of LA and associated oxylipins. LA, linoleic acid. Data points are mean fatty acid and oxylipin concentrations by treatment group and time point and error bars are standard errors. Statistical significance of changes over time within treatment groups and differences between treatment groups in the changes over time is indicated by * P<0.05, ** P<0.01 and *** P<0.0001.

### Omega-6 Fatty Acid Results

#### 3.2.4. AA

Both total blood lipid AA and free AA declined between baseline and 34 weeks’ gestation, with a larger decline seen in the omega-3 LCPUFA group. While AA oxylipin derivatives significantly rose or fell over pregnancy, no differences were observed between the groups **Figure 4**.

#### 3.2.5. LA

There was a small but significant reduction in total blood lipid LA in the control group over the course of pregnancy which was negated by omega-3 LCPUFA treatment. In contrast, free LA increased with gestational age in both groups. However, all LA derived oxylipins decreased over pregnancy independently of the treatment groups **Figure 5**.

Further details, including estimated effects and 95% confidence intervals, can be found in **Supplementary Table 3**. Similar results were obtained in the sensitivity analysis (data not shown).

#### 3.2.6. Effect of baseline omega-3 status on DHA and associated oxylipins

We previously reported that the effects of omega-3 LCPUFA supplementation on early preterm birth could be categorized by omega-3 status at baseline (early pregnancy ∼14 weeks gestation) [12]. For the random cohort reported here, there were 195 (15.4%) women with a baseline omega-3 status of ≤4.1% (low baseline status); 518 (41.0%) >4.1 to ≤4.9% (moderate baseline status), and 550 women (43.5%) >4.9% (high baseline omega-3 status). Baseline omega-3 status significantly influenced the effects of omega-3 LCPUFA supplementation on changes over pregnancy for 7-HDHA and the aggregate of free omega-3 fatty acids (interaction P<0.05), **Table 2**. 7-HDHA showed a decline over pregnancy across the three categories of baseline omega-3 status within the control group. Omega-3 LCPUFA supplementation resulted in no significant change in 7-HDHA over the course of pregnancy for categories ≤4.1% and >4.1% to ≤4.9%, but an increase in women with high baseline status >4.9%, leading to a significant difference between the groups in this baseline category. A similar pattern was also observed for 4-HDHA (interaction P<0.06). Free omega-3 decreased over time in both groups across all three categories of baseline omega-3 status. However, omega-3 LCPUFA supplementation resulted in significantly smaller decreases over time among women with moderate or high omega-3 baseline status, **Table 2**. Additional post hoc comparisons testing for differences between women with high and low baseline status (>4.9% vs ≤4.1%) showed that differences in the change over pregnancy between the groups were significantly larger among women with high compared to low baseline status for free DHA, 4-HDHA, 7-HDHA and the sum of free omega-3 fatty acids, **Supplementary Table 4** and **Supplementary Figures 1 to 4**.

**Table 2:** Effect of baseline omega-3 status, treatment group and time on concentration of DHA, DHA derived oxylipins and omega-3 fatty acids^†^.

| **Baseline Omega-3<sup>†</sup> Status** | **Omega-3 Group Baseline Mean (SD)** | **Control Group Baseline Mean (SD)** | **Omega-3 Group 34 weeks Mean (SD)** | **Control Group 34 weeks Mean (SD)** | **Omega-3 Group Effect\*** | **Control Group Effect\*** | **Difference\*\*** | **Interaction P-value\*\*\*** |
| --- | --- | --- | --- | --- | --- | --- | --- | --- |
| **DHA** |  |  |  |  |  |  |  |  |
| ≤4.1% | 2.03 (0.27) | 1.98 (0.31) | 2.81 (0.91) | 1.96 (0.43) | 0.77 (0.56, 0.99) | -0.02 (-0.11, 0.07) | 0.80 (0.57, 1.03) | 0.19 |
| >4.1 - ≤4.9 | 2.50 (0.33) | 2.52 (0.31) | 3.20 (0.91) | 2.35 (0.42) | 0.71 (0.59, 0.83) | -0.17 (-0.22, -0.11) | 0.87 (0.74, 1.01) |  |
| >4.9% | 3.21 (0.54) | 3.26 (0.61) | 3.64 (0.85) | 2.67 (0.55) | 0.43 (0.31, 0.54) | -0.59 (-0.66, -0.51) | 1.01 (0.88, 1.15) |  |
| **Free DHA** |  |  |  |  |  |  |  |  |
| ≤4.1% | 0.84 (0.59) | 0.72 (0.32) | 0.59 (0.33) | 0.39 (0.16) | -0.26 (-0.38, -0.14) | -0.33 (-0.39, -0.27) | 0.07 (-0.07, 0.20) | 0.06 |
| >4.1 - ≤4.9 | 0.88 (0.48) | 0.88 (0.44) | 0.70 (0.40) | 0.49 (0.20) | -0.17 (-0.25, -0.10) | -0.39 (-0.45, -0.34) | 0.22 (0.13, 0.32) |  |
| >4.9% | 0.90 (0.46) | 0.94 (0.44) | 0.78 (0.40) | 0.56 (0.25) | -0.12 (-0.19, -0.05) | -0.38 (-0.44, -0.33) | 0.26 (0.17, 0.35) |  |
| **4-HDHA** |  |  |  |  |  |  |  |  |
| ≤4.1% | 20.04 (29.41) | 13.33 (11.48) | 10.35 (8.48) | 7.41 (5.43) | -9.52 (-15.49, -3.54) | -5.93 (-8.19, -3.67) | -3.59 (-9.98, 2.80) | 0.06 |
| >4.1 - ≤4.9 | 18.79 (24.32) | 17.88 (18.95) | 13.73 (12.60) | 9.44 (6.63) | -5.13 (-8.49, -1.77) | -8.47 (-10.95, -5.98) | 3.34 (-0.85, 7.52) |  |
| >4.9% | 16.59 (14.23) | 17.89 (14.31) | 14.52 (13.58) | 10.76 (8.13) | -1.96 (-4.42, 0.49) | -7.05 (-9.01, -5.09) | 5.08 (1.93, 8.23) |  |
| **7-HDHA** |  |  |  |  |  |  |  |  |
| ≤4.1% | 0.96 (1.13) | 0.85 (0.64) | 0.83 (0.62) | 0.59 (0.29) | -0.13 (-0.32, 0.06) | -0.26 (-0.38, -0.15) | 0.13 (-0.09, 0.35) | 0.02 |
| >4.1 - ≤4.9 | 1.13 (1.83) | 0.97 (0.80) | 1.15 (1.04) | 0.75 (0.39) | 0.01 (-0.25, 0.27) | -0.23 (-0.33, -0.12) | 0.24 (-0.04, 0.52) |  |
| >4.9% | 0.95 (0.67) | 1.05 (0.68) | 1.29 (0.97) | 0.86 (0.46) | 0.33 (0.19, 0.48) | -0.19 (-0.28, -0.09) | 0.52 (0.35, 0.69) |  |
| **14-HDHA** |  |  |  |  |  |  |  |  |
| ≤4.1% | 6.18 (8.72) | 5.65 (6.79) | 5.51 (8.02) | 3.16 (3.29) | -0.42 (-2.67, 1.84) | -2.53 (-4.03, -1.02) | 2.11 (-0.61, 4.82) | 0.27 |
| >4.1 - ≤4.9 |  |  |  |  |  |  |  |  |

| Baseline Omega-3<sup>†</sup> Status | Omega-3 Group Baseline Mean (SD) | Control Group Baseline Mean (SD) | Omega-3 Group 34 weeks Mean (SD) | Control Group 34 weeks Mean (SD) | Omega-3 Group Effect* | Control Group Effect* | Difference** | Interaction P-value*** |
| --- | --- | --- | --- | --- | --- | --- | --- | --- |
| >4.9% | 7.35 (5.98) | 9.38 (10.46) | 6.20 (5.39) | 4.10 (3.37) | -1.13 (-2.11, -0.14) | -5.23 (-6.47, -4.00) | 4.11 (2.53, 5.69) |  |
| **19,20EpDPA** |  |  |  |  |  |  |  |  |
| ≤4.1% | 0.24 (0.30) | 0.18 (0.27) | 0.31 (0.25) | 0.19 (0.18) | 0.05 (-0.01, 0.12) | -0.00 (-0.05, 0.05) | 0.05 (-0.03, 0.14) | 0.14 |
| >4.1 - ≤4.9 | 0.25 (0.31) | 0.26 (0.32) | 0.45 (0.35) | 0.30 (0.21) | 0.18 (0.13, 0.24) | 0.02 (-0.02, 0.07) | 0.16 (0.09, 0.23) |  |
| >4.9% | 0.33 (0.42) | 0.32 (0.33) | 0.50 (0.39) | 0.44 (1.49) | 0.15 (0.09, 0.22) | 0.11 (-0.09, 0.30) | 0.04 (-0.16, 0.25) |  |
| **Omega-3<sup>†</sup> fatty acids** |  |  |  |  |  |  |  |  |
| ≤4.1% | 3.76 (0.34) | 3.70 (0.36) | 4.49 (1.06) | 3.53 (0.85) | 0.73 (0.47, 0.99) | -0.17 (-0.35, 0.01) | 0.90 (0.59, 1.21) | 0.15 |
| >4.1 - ≤4.9 | 4.52 (0.23) | 4.53 (0.22) | 5.03 (1.17) | 4.09 (0.74) | 0.51 (0.35, 0.67) | -0.45 (-0.55, -0.35) | 0.96 (0.77, 1.14) |  |
| >4.9% | 5.63 (0.70) | 5.71 (0.79) | 5.59 (1.10) | 4.48 (0.74) | -0.04 (-0.20, 0.11) | -1.23 (-1.35, -1.12) | 1.19 (0.99, 1.38) |  |
| **Free Omega-3<sup>†</sup> fatty acids** |  |  |  |  |  |  |  |  |
| ≤4.1% | 1.04 (0.73) | 0.88 (0.39) | 0.72 (0.39) | 0.52 (0.21) | -0.31 (-0.46, -0.17) | -0.37 (-0.44, -0.29) | 0.05 (-0.11, 0.21) | 0.04 |
| >4.1 - ≤4.9 | 1.08 (0.60) | 1.09 (0.54) | 0.89 (0.51) | 0.63 (0.26) | -0.19 (-0.29, -0.10) | -0.45 (-0.52, -0.38) | 0.26 (0.14, 0.38) |  |
| >4.9% | 1.10 (0.55) | 1.15 (0.52) | 0.97 (0.50) | 0.73 (0.32) | -0.13 (-0.21, -0.04) | -0.42 (-0.49, -0.36) | 0.30 (0.19, 0.40) |  |

DHA, docosahexaenoic acid;
<sup>†</sup>Omega-3, sum of docosahexaenoic acid, eicosapentaenoic acid, docosapentaenoic acid and alpha linolenic acid.
\* Values are estimated difference in the mean fatty acid or oxylipin concentrations (95% confidence interval) for 34 weeks' vs baseline adjusted for enrolment centre and recent omega-3 supplementation by baseline total omega-3 status. Positive values indicate fatty acid or oxylipin concentrations are increasing over time. Negative values indicate fatty acid or oxylipin concentrations are decreasing over time.
\*\* Values are estimated difference in the mean change in fatty acid or oxylipin concentrations over time (95% confidence interval) for omega-3 vs control adjusted for enrolment centre and recent omega-3 supplementation by baseline total omega-3 status.
\*\*\* P-value for the 3-way interaction between baseline omega-3 status, treatment group and time. Results should be interpreted with caution where the interaction is not statistically significant.

## 4. Discussion and Conclusions

We anticipated that omega-3 LCPUFA supplementation would mostly affect DHA and DHA-derived oxylipins rather than EPA and EPA-derived oxylipins, because of the small dose of EPA included in the study intervention. We also assumed that supplementation of women with an oil blend rich in DHA would increase the levels of DHA and its metabolites in dried blood spots. Thus, the increased total blood lipid DHA, 7-HDHA, and 19,20 EpDPA due to supplementation seemed logical. We had not anticipated the fact that several of the oxygenated metabolites of DHA naturally declined over the assessed 20-week period of gestation. However, omega-3 LCPUFA supplementation reduced the declines of free DHA, 4-HDHA, and 14-HDHA between the end of the first trimester of pregnancy and 34 weeks of gestation so that overall the effect of supplementation was consistent in that there was an overall increase in DHA and its various metabolites relative to no supplementation.

Although early preterm births would have occurred between the two sampling periods in our study (14 and 34 weeks of gestation) we nevertheless considered it important to understand these biochemical data in the context of the clinical outcomes from the ORIP trial. Interestingly, secondary analyses of the ORIP Trial showed that the benefit of omega-3 LCPUFA supplementation on reducing the risk of early preterm birth was limited to women with low baseline omega-3 status (≤4.1%); women who already had a higher baseline status of omega-3 fatty acids (>4.9%) and who were supplemented with additional DHA, increased their risk of early preterm birth [12]. Thus, it is pertinent to explore the relationships between changes in oxylipins recorded here according to these measures of omega-3 status. In particular, whether the changes in oxylipins in relation to baseline measures and effects of omega-3 LCPUFA supplementation could explain the apparently contradictory effects of supplementation on clinical outcomes. Namely, why omega-3 LCPUFA supplementation lowered the risk of early preterm birth in women with low omega-3 status at baseline, while the same treatment increased the risk of early preterm birth in women with high omega-3 status at baseline. Our data demonstrated that it was the 5-lipoxygenase derivatives, 7-HDHA and 4-HDHA, whose response to omega-3 LCPUFA supplementation was influenced by the baseline omega-3 status. Omega-3 LCPUFA supplementation in women with higher omega-3 status at baseline was associated with a significant increase in 7-HDHA and 4-HDHA between treatment and control whereas there were no differences between groups in 7-HDHA and 4-HDHA in women with moderate or low omega-3 status. It is therefore feasible that the increase in the 5-lipoxygenase derivatives associated with DHA supplementation in women with already high omega-3 fatty acid status may be causally related to the increased risk of early preterm birth in these women. However, there was no evidence of 7-HDHA and 4-HDHA being affected by omega-3 LCPUFA supplementation in women with low omega-3 status, where risk of early preterm birth was high, and supplementation effectively lowered this risk. Further exploration is needed to understand the role of 5-lipoxygenase products as well as other oxylipins in in determining the timing of birth at both high and low baseline omega-3 status.

The changes seen in the current report were consistent with a much smaller study also nested within the ORIP trial that involved plasma samples (rather than whole blood from dried blood spots) collected at trial entry (end of the first trimester) and 24 weeks of gestation, indicating that the observed changes happen relatively quickly [21]. The clinical significance of these changes is yet to be fully determined, although this earlier study [21] did hypothesize that higher 5-lipoxygenase derivatives of both DHA and AA in early to mid-pregnancy were associated with increased risk of spontaneous preterm birth.

Traditionally, the timing of parturition has been discussed in terms of cyclooxygenase products, largely involving a preponderance of the AA derivatives PGE2 and PGF2α, with the equivalent derivatives from EPA being significantly less potent [9, 22, 23]. Our technology was not sensitive enough to accurately detect these cyclooxygenase products in the collected dried blood spots, although the pattern of change of a related prostaglandin (PGD2) across pregnancy did not differ between the omega-3 LCPUFA and control groups.

Surprisingly, our data showed very little, if any, difference in the pattern of change of total lipid and free omega-6 AA and LA between omega-3 supplemented and control groups, and this lack of difference also resulted in no differences between groups in their measured oxylipin derivatives. In non-pregnant adults and lactating women, the equivalent omega-3 LCPUFA supplementation with 800mg DHA would be expected to reduce blood AA levels by more than the small reduction observed in this study [24]. It may be that the hormonal and hemodynamic changes that occur over the course of pregnancy are partially designed to maintain an AA-associated homeostasis. Interestingly, from the oxylipins assessed it was only three AA derived oxylipins, which are 5-lipoxygenase products (5-HETE, LXA4, LXB4), that naturally increased over the course of pregnancy regardless of omega-3 supplementation..

Our study had several strengths. It was nested in a randomized controlled trial, had large numbers and matched fatty acids in total blood lipids with the free fatty acid pool and their derived oxylipins, providing us with some important insights into how fatty acids and oxylipins change over the course of pregnancy, and how this trajectory is influenced by omega-3 LCPUFA supplementation. A study such as this would have been more difficult logistically and more costly without the use of the dried blood spot technology, although we were not able to consistently detect the cyclooxygenase products. The main limitation is that we were not able to reliably link the fatty acid and oxylipin changes between 14 and 34 weeks of gestation directly with prematurity, as we would have missed all the preterm births occurring before 34 weeks of gestation.

In summary, this is a large and comprehensive study to document fatty acid and oxylipin changes across pregnancy. We demonstrated that most fatty acids in both total blood lipid and free fatty acid fractions and their associated oxylipins decline over the course of pregnancy, with a few notable exceptions related to 5-lipoxygenase products. Omega-3 LCPUFA supplementation mostly arrested the declines in free DHA and DHA-derived oxylipins, with a suggestion of a differential response in the 5-lipoxygenase products depending on the baseline omega-3 status of women. Our work has added to the speculation that 5-lipoxygenase products have an important role to play in modulating pregnancy duration. Further work to understand their role will allow us to better tailor treatments for women to achieve optimal pregnancy outcomes.

## Data Availability

Data will be available to Researchers who provide a methodologically sound research proposal following review and approval by the trial steering committee and completion of a signed data access agreement.
Following approval, de-identified, individual participant data that underlie the results reported in this article (text, tables, figures and appendices) and/or dataset(s) will be limited to those participants and variables that are necessary for completion of the approved research proposal.
Data sharing requests for de-identified raw data should be made to the trial steering committee and can be submitted to

## Abbreviations used

AA: arachidonic acid
ALA: alpha linolenic acid
DBS: dried blood spot
DHA: docosahexaenoic acid
DPA: docosapentaenoic acid
EPA: eicosapentaenoic acid
FAME: fatty acid methyl esters
LA: linolenic acid
LC-MS-MS: Liquid Chromatography with tandem mass spectrometry
LCPUFA: long chain polyunsaturated fatty acid
ORIP: Omega-3 fats to Reduce the Incidence of Prematurity
PUFA: polyunsaturated fatty acids.

## Acknowledgements

We thank the women and their families for their participation in the original study.

## Contribution to Authorship

RAG and MM, conceived the analysis. KPB, RAG, LNY, JG and MM were involved in the analysis and interpretation of data and SL and LNY carried out the statistical analyses. KB drafted the manuscript and all authors critically revised and edited the manuscript and approved the final submitted version. All authors take responsibility for the integrity of the data and the accuracy of the data analysis.

